# Rethinking covariate adjustment in psychiatric biomarker research: a framework applied to UK Biobank blood samples

**DOI:** 10.64898/2026.04.19.26351233

**Authors:** Mirim Shin, Jacob J Crouse, Ian B Hickie, Naomi R Wray, Clara Albiñana

## Abstract

**Importance:** Blood-based biomarkers hold promise for psychiatric diagnosis and prognosis, yet clinical translation is constrained by poor reproducibility. Psychiatric biomarker studies are typically small, and demographic, behavioral, and temporal covariates often go undetected or cannot be adequately modeled. This may lead to residual confounding and unstable associations.

**Observations:** Leveraging UK Biobank data (N=~500,000), we systematically quantified how technical, demographic, behavioral, and temporal covariates influence 29 blood biomarkers commonly measured in research studies in psychiatry. Variance analyses showed substantial differences across biomarkers. Technical factors explained 1-6% and demographic factors explained 5-15% of the variance, with pronounced age-by-sex interactions for lipids and sex hormones. Behavioral covariates, particularly body mass index (BMI) and smoking, strongly influenced inflammatory markers. Temporal factors introduced systematic confounding. Chronotype was associated with blood collection time, multiple biomarkers exhibited marked diurnal rhythms (including testosterone, triglycerides, and immune markers), and inflammatory markers showed seasonal peaks in winter. In association analysis of biomarkers with major depression, bipolar disorder and schizophrenia, covariate adjustments attenuated or eliminated a substantial proportion of the biomarker-disorder associations, with BMI emerging as the dominant confounder. These findings demonstrate that such confounding structures exist and can be characterized in large cohorts, though specific biomarker-disorder relationships require validation in clinical samples.

**Conclusions and Relevance:** Poor reproducibility of biomarkers may not only stem from insufficient biological signal but also from inconsistent handling of confounders. We propose a systematic framework distinguishing technical factors (to be removed), demographic factors (addressed through adjustment or stratification), temporal factors (ideally controlled at design stages), and behavioral factors (requiring explicit causal reasoning). Associations robust to multiple adjustment strategies should be prioritized for clinical biomarker development. Standardized collection protocols, comprehensive covariate measurement, and transparent reporting across models are essential to improve reproducibility and identify biomarkers that reflect genuine illness-related pathophysiology.

## INTRODUCTION

Blood-based biomarkers are increasingly studied with goals to improve psychiatric diagnosis, prognosis, treatment selection, and prediction of treatment response.^1^ For example, inflammatory markers such as leukocytes, haptoglobin, and C-reactive protein (CRP) are associated with psychiatric disorder risk before diagnosis,^2^ and inflammation has been proposed for identifying a specific clinical phenotype of major depression in the future DSM-6.^3^ These findings highlight the potential of blood biomarkers for early risk detection and mechanistic insight.

Despite this, the field lacks robust, reliable, and valid psychiatric biomarkers. A fundamental obstacle to clinical translation is confounding and methodological heterogeneity, which compromise reproducibility.^4^ Many candidate biomarkers fail validation because they reflect nonspecific influences (e.g., demographics, lifestyle, treatment effects, or illness chronicity) rather than disorder-specific pathophysiology.^5^

Several key challenges have emerged. First, the transdiagnostic nature of most peripheral biomarkers (e.g., brain-derived neurotrophic factor [BDNF], CRP, interleukins [ILs]) suggests that they might reflect systemic consequences of psychiatric disorders rather than disorder-specific etiology, thereby, necessitating precise covariate control.^1^ Second, biomarker-symptom associations are often strongly attenuated or become null when demographic, lifestyle (e.g., body mass index [BMI], smoking), or chronic disease-related covariates are accounted for.^6^ Third, technical variation is high, with measurement quality critically dependent on biofluid type, venipuncture-to-freezing time, centrifugation procedures, storage temperature, freeze-thaw cycles, and assay methodology.^5^ In a UK Biobank [UKB] analysis of 35 biomarkers, covariate-explained variance ranged from 1.7% to 90%, with different variables contributing uniquely to each.^7^

Failure to address these variables appropriately increases the risk of spurious findings.^1^ However, determining appropriate covariate adjustment strategies is complex. Some variables may function as true confounders (warranting adjustment) or as mediators (for which adjustment may remove meaningful illness-related signals), and their treatment depends on the research question and study design. Temporal factors illustrate this ambiguity. Time of day at sample collection, rarely reported in psychiatric biomarker studies, may act as a confounder (when independent of case-control status) or a mediator (when chronotype-driven scheduling preferences influence appointment timing). Similarly, although depressive symptoms follow seasonal patterns, season of assessment also introduces environmental variation (e.g. viral infections, allergens, weather, daylight exposure) that independently affects biomarker levels. Because many biomarkers follow circadian and seasonal rhythms, this variability is best minimized through standardized collection protocols at the study design stage rather than through post-hoc statistical adjustments.

Psychiatric biomarker studies are further limited by small sample sizes (typically n=50-200), which are insufficient to support the complex covariate models needed to disentangle these effects. Confounding structures are likely to remain undetected.

To address some of these issues, we leveraged UKB data to develop a systematic empirical framework for covariate adjustment in psychiatric biomarker research. The large sample size of UKB enables characterization of confounding structures at a scale not achievable in smaller cohorts. We selected 29 blood biomarkers commonly used in psychiatric research across four analytical platforms: biochemistry, blood cell counts, metabolomics-NMR, and proteomics-Olink. These biomarkers have been previously characterized in UKB,^7^ but our focus is to evaluate them specifically from the perspective of psychiatric research and to develop a framework for covariate adjustment. Our objectives were to: (1) quantify the variance explained in each biomarker by technical, demographic, temporal, and behavioral variables; (2) quantify associations between these variables and psychiatric disorders; (3) evaluate how covariate adjustment alters biomarker-disorder associations; and (4) develop an empirical framework for determining appropriate adjustment strategies for different covariate types.

## METHODS

### Study population and data source

This study utilized data from UKB, a prospective cohort of ~500,000 participants aged 40-69 years recruited between 2007 and 2010.^8^ This analysis was conducted under application number 116122.

### Blood biomarker selection and platforms

Through a targeted literature review of blood-based markers that are frequently reported in psychiatric research and are available in the UKB (see Table S1), we selected 29 biomarkers across four analytical platforms: biochemistry (11 markers), blood cell counts (4 markers), NMR metabolomics (4 markers), and Olink proteomics (10 markers). The full biomarker list and platform-specific protocols are detailed in eMethods. All biomarkers were filtered to exclude participants having fasting time ≥24 hours, sample collection time outside 9:00-20:00 hours and with categorical variables n<50.

### Covariate classification

Covariates were classified into four categories: (1) *Technical factors*: assessment center, genetic principal components (PC1-PC20), fasting time, and platform-specific variables (e.g., batch, device, processing time); (2) *Demographic factors*: age, sex, and age-by-sex interactions; (3) *Temporal factors*: time of day at blood collection and month of assessment; (4) *Behavioral factors*: BMI, smoking status, and chronotype. Although genetic PCs capture ancestry variation, they are treated here as technical factors requiring universal adjustment; biomarker levels were residualized for all technical covariates prior to downstream analyses. Ancestry was additionally examined as a demographic factor in covariate-disorder association analyses. Full covariate definitions and UKB field identifiers are provided in eMethods.

### Psychiatric disorder definitions

Participants were classified as cases based on ICD-9/10 codes from hospital in-patient records, death registers, and self-reported diagnoses for: bipolar disorder (BD; F31), major depressive episode (MDE; F32), recurrent depression (RD; F33), and schizophrenia (SCZ; F20). See eMethods for full field identifiers.

### Statistical analysis

#### Variance quantification

We quantified variance explained by each covariate using sequential linear models, with partial R^2^ values calculated as the ratio of each covariate term’s sum of squares to total sum of squares.

#### Covariate-psychiatric disorder associations

We tested associations between key covariates (sex, age, season, smoking, BMI, chronotype, time of day, and ancestry) and psychiatric diagnoses using logistic regression. FDR-corrected p-values (*P*FDR) are reported; associations with *P*FDR<.05 are considered significant.

#### Sequential adjustment analysis

To evaluate the effects of relevant covariate on biomarker-disorder associations, we first adjusted all biomarker levels for technical covariates in a linear model (biomarker ~ technical factors) and took the model’s residuals. We then tested associations between each residualized biomarker and psychiatric disorders under multiple adjustment levels: (1) unadjusted; (2) adjusted for individual covariates (time of day, age, sex, chronotype, season, smoking, BMI) separately; and (3) fully adjusted (all covariates jointly). We compared the proportion of significant associations retained across adjustment levels. Details on temporal factor analyses, chronotype x study design comparisons, and statistical model specifications are provided in eMethods.

## RESULTS

Demographic and behavioral characteristics of the UKB study sample are presented in Table S2. Cases showed higher rates of current smoking, higher BMI, and greater evening chronotype preference across all disorder categories.

### Sources of biomarker variability

The proportion of variance explained by covariates varied substantially across biomarkers, ranging from near zero for IL-10 to more than 90% for testosterone (Figure S1, Data S2). Sex was the dominant factor for sex hormones, explaining 90% of variance in testosterone and 23% in estradiol, but also contributed substantially to non-hormonal markers (e.g., 27% leptin, 33% creatinine). Age explained 12% of variance for cystatin C and 6% for HbA1c, noting that UKB participants are all aged over 40 years. Age x sex interactions are presented in Figure S3. BMI accounted for substantial variance in metabolic and inflammatory markers, including 40% for leptin, 18% for CRP and 12% for IL-6. Technical factors explained up to 6% of variance overall; further examination of technical variables (Figure S2 and Data S2) indicated substantial contributions across multiple biomarkers. Genetic ancestry was associated with unadjusted biomarker levels across multiple biomarkers (Figure S3); but these differences were fully attenuated after adjustment for genetic PCs (Figure S4).

Temporal factors explained a small proportion of overall biomarker variance (Figure S1) but showed substantial diurnal and seasonal effects for specific biomarkers (Figure 1). Time of blood collection had pronounced effects: testosterone declined sharply from morning to mid-afternoon, triglycerides peaked mid-afternoon with lower levels in both morning and evening, and alanine, BDNF, IL-1β, and IL-10 declined progressively from morning to evening, whereas IL-6 and leukocyte count increased throughout the day. Seasonally, inflammatory markers (CRP, IL-6, leukocyte count) peaked in winter and were lowest in summer; HbA1c and IL-1β showed sinusoidal patterns peaking mid-year; IL-18 and total cholesterol showed bimodal distributions.

**Figure 1.**
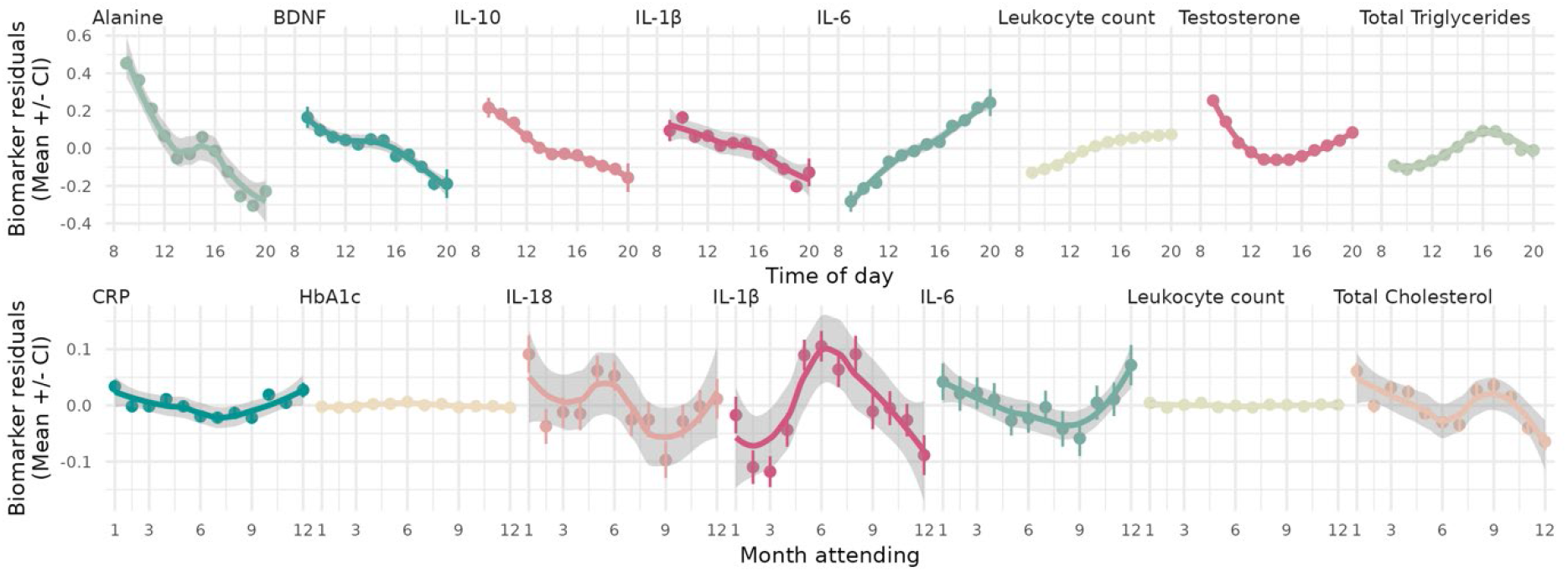
Diurnal and seasonal and variation in selected biomarker residuals across time of day and calendar months. **Note:** Biomarkers shown are those with significant diurnal or seasonal variation out of the 29 studied, after adjustment for technical covariates. Dots represent mean residual values per hour (top) or per month (bottom); error bars show 95% confidence intervals; lines show loess smoothing function.

Chronotype was associated with blood collection time, most strongly when participants had scheduling flexibility. We compared the baseline visit (UKB1; assigned appointments) and the first repeat visit (UKB2; self-scheduled; see eMethods). Among 6,690 participants attending both visits, individuals reporting a “definitely evening” chronotype had blood drawn 58 minutes later than those reporting “definitely morning” chronotype in UKB2 (14:19 vs 13:21), compared to only a 15-minute difference in UKB1 (14:15 vs 14:00; interaction β=0.723, *P*=1.23×10^-4^). These findings indicate that chronotype contributes systematic, rather than random, variation in blood collection timing across UKB visits (Figure S6).

### Covariate associations with psychiatric disorders

Covariates were significantly associated with psychiatric diagnoses (Figure 2, Data S3). Male sex was associated with lower odds of MDE and RD but higher odds of SCZ. Older age was associated with lower odds of all diagnoses, potentially reflecting survivor bias. Non-European ancestry groups showed differential associations with psychiatric diagnoses, particularly African ancestry with SCZ. Later appointment times (after 12:00) and a “definitely evening” chronotype were associated with increased odds across all four disorders. Higher BMI was associated with MDE and RD, with a characteristic U-shape relationship. Current smoking status was also associated with elevated odds of all disorders, particularly SCZ.

**Figure 2.**
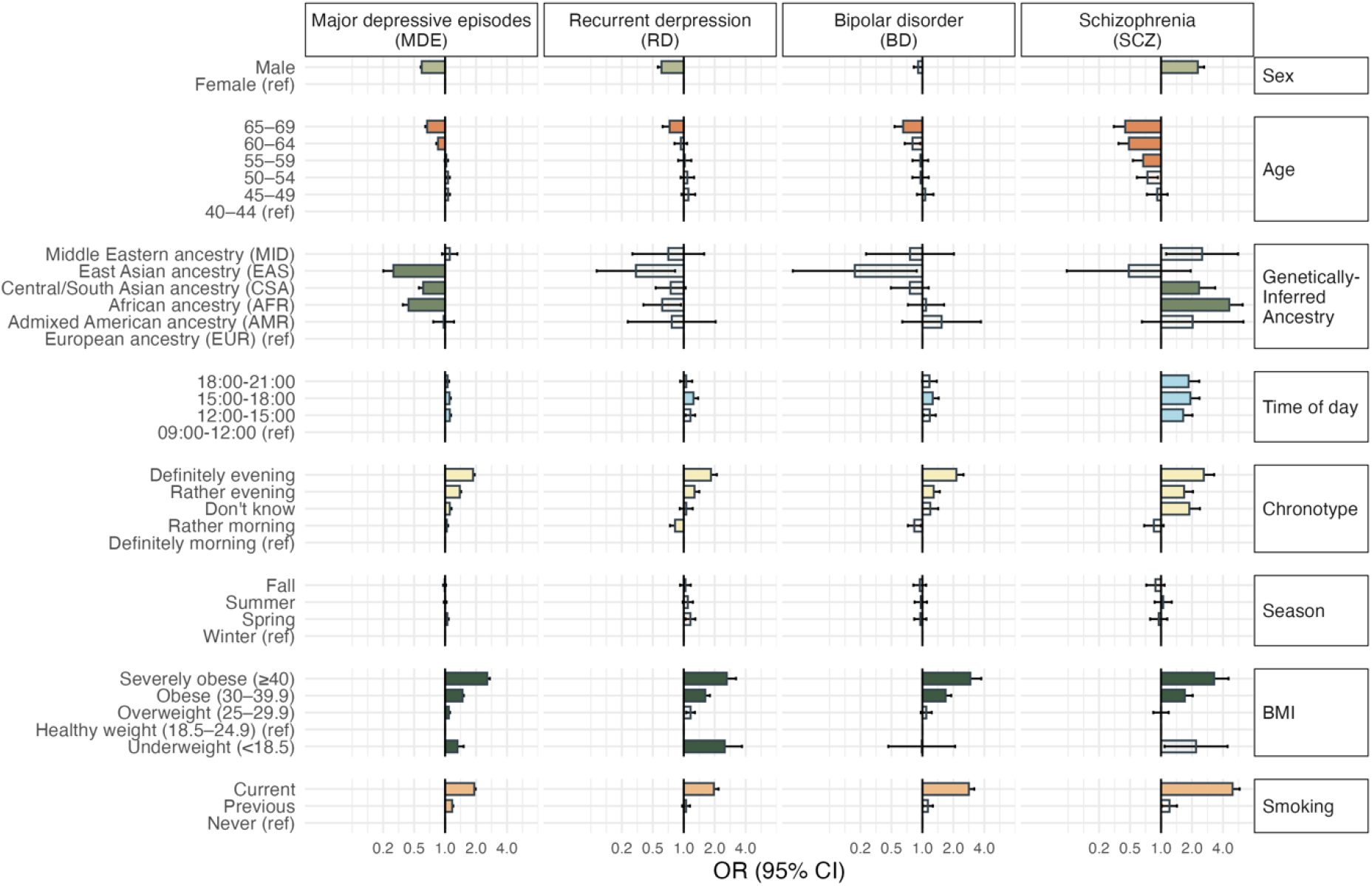
Associations between demographic, behavioral, and temporal covariates and psychiatric disorder diagnoses. **Note:** FDR-significant associations are indicated with colored bars, with different colors used to distinguish the y-axis categories.

### Impact of covariate adjustment on biomarker-disorder associations

Of the 29 biomarkers examined, 26 (excluding alanine, BDNF and IL-1β) were associated with at least one disorder in unadjusted models (Figure 3&S7, Data S4). The proportion of associations surviving full adjustment varied by disorder. BD retained 9 of 13 associations, SCZ 10 of 14, MDE 13 of 19, while RD showed the greatest attenuation (3 of 12), reflecting its smaller case sample (~2,600 for RD vs ~40,000 for MDE).

**Figure 3.**
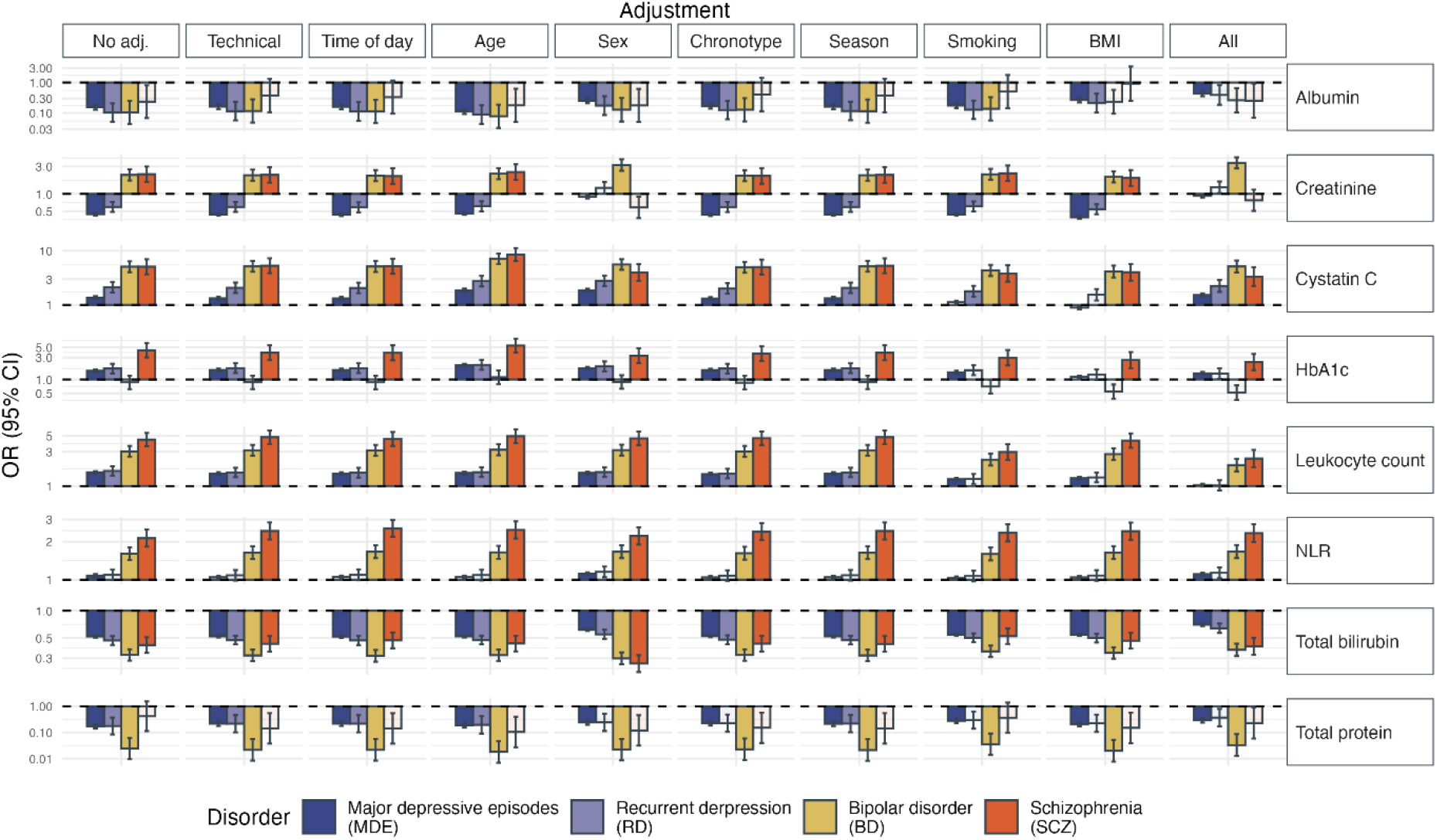
Top biomarker-disorder associations with large effect sizes (OR >2 or < 0.5) under sequential covariate adjustment. **Note:** Bars show odds ratios for each biomarker-disorder association. Columns represent adjustment levels: No adj. (unadjusted), Technical (technical covariates only), individual covariates (time of day, age, sex, chronotype, season, smoking, BMI), and All (fully adjusted). Ancestry variation is controlled via residualization for genetic principal components (see Methods). Individual covariate adjustments were performed on biomarker residuals after technical adjustment. Figure S7 shows all 29 biomarkers.

In the fully adjusted models, cystatin C and total bilirubin were associated with all four disorders. Cystatin C showed particularly large effect sizes: BD (OR, 5.13; *P*FDR=3.93×10^-33^), SCZ (OR, 3.34; *P*FDR=2.29×10^-6^), RD (OR, 2.24; *P*FDR=2.84×10^-7^), MDE (OR, 1.52; *P*FDR=1.20×10^-26^). Total bilirubin showed inverse associations, strongest for BD (OR, 0.37; *P*FDR=1.62×10^-32^) and SCZ (OR, 0.41; *P*FDR=1.83×10^-13^). Other notable associations included leukocyte count and Neutrophil-to-Lymphocyte Ratio (NLR) with BD and SCZ, and total protein with BD (OR, 0.03; *P*=2.65×10^-9^) and MDE (OR, 0.29; *P*FDR=5.50×10^-30^). Full results are provided in Data S4.

Sequential adjustment for individual covariates showed distinct confounding patterns. BMI was the most influential confounder overall. For BD, BMI adjustment attenuated associations with IL-6, IL-12, albumin, and CRP; for MDE, with cystatin C, HbA1c, IL-12, IL-6, and TNFα; for RD, with cystatin C, HbA1c, total protein, total triglycerides, leukocyte count, and Monocyte-to-Lymphocyte Ratio (MLR). Sex was particularly influential for SCZ. Smoking was a major confounder for RD (HbA1c, total protein, total triglycerides, leukocyte count, MLR), with smaller effects for BD (Gamma-Glutamyl Transferase [GGT], IL-6) and MDE (cystatin C, GGT). Chronotype, time of day, and season had limited confounding effects (Figure 3&S7, Data S4).

To determine whether associations were driven by general inflammatory processes, we evaluated additional adjustment for leukocyte count. Although most biomarkers correlated with leukocyte count (Figure S8A), further adjustment did not substantially alter the pattern of associations (Figure S8B), suggesting the observed effects are not solely attributable to variation in white blood cell levels.

## DISCUSSION

We selected 29 blood-based biomarkers in UKB to characterize covariate contributions and propose a framework for appropriate adjustment in psychiatric biomarker research. Importantly, UKB enables systematic characterization of confounding structures at a scale not achievable in smaller cohorts, akin to the large-scale epidemiological studies that showed striking biomarker-disorder associations undetectable at moderate sample sizes.^9^ We show that many commonly studied biomarkers are substantially influenced by demographic, behavioral, temporal, and technical factors that are often inadequately handled in psychiatric studies because of limited sample size or incomplete covariate measurement. A substantial proportion of biomarker-disorder associations were attenuated or eliminated following covariate adjustment, with BMI and smoking emerging as particularly influential confounders. Poor reproducibility may therefore stem from inconsistent covariate handling rather than insufficient biological signal alone.

Impact of covariates varied substantially across biomarkers, from near zero (IL-10) to over 90% (testosterone), indicating that one-size-fits-all adjustment strategies are inappropriate. Some biomarkers (albumin, total protein) showed minimal variance explained by measured covariates, possibly reflecting tight homeostatic regulation,^10^ whereas sex hormones, leptin and inflammatory markers were highly susceptible to demographic and behavioral influences.

Age-by-sex interactions extended beyond sex hormones, including the well-documented lipid crossover at menopause, where women surpass men in LDL cholesterol after age 50-55.^11,12^ These interactions highlight the limitations of additive adjustment for “age + sex” without interaction terms or stratification, which may result in residual confounding and biased estimates. Adjustment for genetic PCs removed ancestry-related biomarker differences (Figures S3-S4), consistent with standard practice to control population stratification in genetics studies but rarely included in biomarker studies. Importantly, substantial ancestry-associated differences in biomarker levels persist before PC adjustment (Figure S3), and ancestry is independently associated with psychiatric diagnoses (Figure 2). In studies without genomic data, self-reported ancestry should therefore be treated as an essential demographic covariate rather than omitted.

Temporal factors introduced systematic rather than random confounding. Inflammatory markers (CRP, IL-6, leukocytes) peaked in winter, likely reflecting seasonal infectious disease patterns,^13^ whereas HbA1c peaked mid-year, potentially reflecting seasonal metabolic changes.^14^ Diurnal patterns were also pronounced, with testosterone declining from morning to afternoon, and immune markers increasing throughout the day, reflecting circadian system’s regulation of metabolic and immune processes.^15,16^ Chronotype was associated with blood collection time—an effect amplified when participants had scheduling flexibility—creating systematic sampling variation that can generate spurious case–control differences.

BMI emerged as the most influential confounder, consistent with adipose tissue as a source of inflammatory markers such as IL-6 and CRP.^17^ Adjustment for BMI eliminated several associations, including IL-6 with BD, RD and SCZ, replicating previous findings.^6^ Both BMI and smoking present interpretational challenges in cross-sectional data. The causal relationship between smoking and psychotic conditions has been suggested by Mendelian randomization.^18^ In addition to having a causal role, they may be confounders (predating onset), mediators (consequences of illness/treatment), or colliders (sharing common causes) depending on temporal relationships with illness onset (Figure 4). Distinguishing these roles requires longitudinal designs that track biomarkers before and after disorder onset, as prospective studies suggest inflammatory markers can both precede psychiatric diagnoses by decades^2^ and follow them as consequences of illness or treatment.

**Figure 4.**
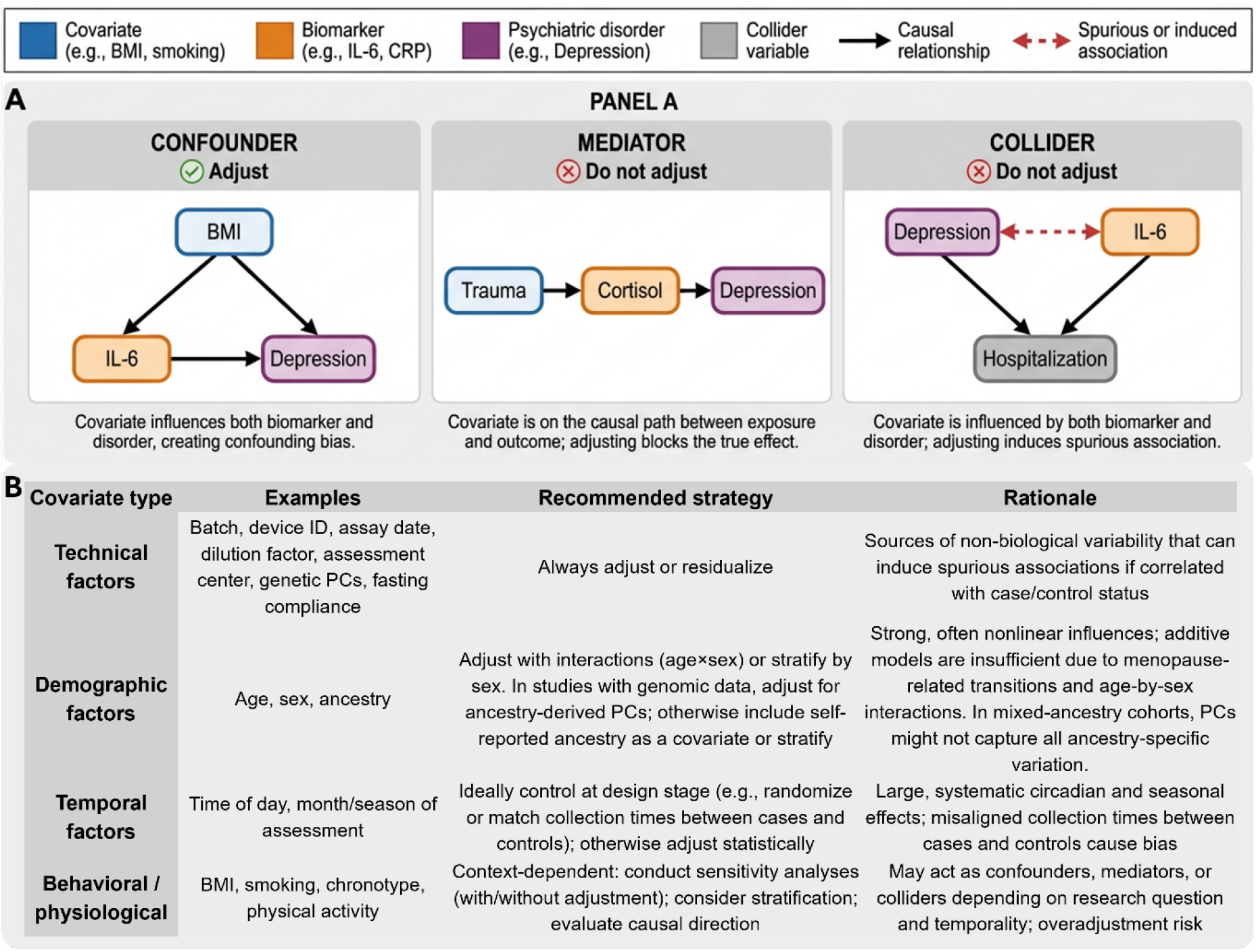
Causal structures and framework for covariate handling in psychiatric biomarker research. **Note:** “Adjust” means fit as covariates in the model. “Residualize” means regress the biomarker on covariates and use the residuals as the outcome variable. In smaller studies where full covariate adjustment is not feasible, these considerations should be addressed at the design stage—for example, by matching cases and controls on key confounders. When validating findings across cohorts, it is particularly important to ensure that confounders present in the discovery cohort are not systematically replicated in the validation cohort.

Several associations persisted after full adjustment. Cystatin C showed strong positive associations across all four diagnoses. Although primarily a renal marker, cystatin C is also influenced by inflammation, thyroid dysfunction, and corticosteroid use.^19^ Total bilirubin showed inverse associations across diagnoses, particularly for bipolar disorder and schizophrenia. The existing literature is heterogeneous—a meta-analysis of 18 studies in treatment-naïve first-episode patients found no significant overall difference from controls,^20^ and the direction of medication effects on bilirubin does not readily explain our findings.^21^ To our knowledge, this is the first transdiagnostic characterization of inverse bilirubin associations at the scale of the UK Biobank and warrants dedicated replication. The transdiagnostic pattern suggests shared pathophysiology or general illness burden rather than disorder-specific mechanisms,^1^ and was not attributable to leukocyte count variation (Figure S8).

Given publication bias favoring positive findings and common practice of reporting minimally adjusted models, some published biomarker-disorder associations may reflect residual confounding rather than disorder-specific pathophysiology. Notably, attenuation reflected reductions in effect size rather than merely loss of statistical significance, support true confounding rather than power artifacts. Associations that remain robust across adjustment strategies should therefore be prioritized for mechanistic follow-up and clinical biomarker development, including proposals such as inflammation-based subtyping of depression that require evidence of persistence after comprehensive adjustment.^3^

Our findings support a structured approach to covariate handling, rather than minimal adjustment (risking confounding) or indiscriminate “kitchen sink” models (risking removal of true biological signal; a concern formalized in the genetics literature, where adjusting for mediators or independent risk factors in case-control designs can reduce true signal or power^22^). We propose a framework (Figure 4) distinguishing technical factors (requiring systematic control), demographic factors (requiring adjustment with interactions or stratification), temporal factors (best addressed through standardized collection procedures), and behavioral factors (requiring careful causal reasoning to distinguish confounders from mediators). This framework emphasizes deliberate measurement, transparent reporting across adjustment models, and alignment of covariate strategy with study design and research questions. Deeply phenotyped, harmonized cohorts—as in the Accelerating Medicines Partnership Schizophrenia program^23^—can similarly show how unmodeled covariate structure reshapes biomarker-disorder associations. When designing validation studies, it is particularly important to ensure confounders present in the discovery cohort (such as smoking, BMI, and genetic ancestry) are not replicated in the validation cohort.

Several limitations of this study warrant consideration. First, UKB participants were aged 40-70 years, limiting generalizability to younger populations in whom different confounding patterns may emerge (due to developmental, hormonal or illness course factors). Second, psychiatric diagnoses relied on hospital inpatient records and self-report, potentially missing undiagnosed cases.^24^ Participants were unlikely to be acutely unwell at assessment, and UKB psychiatric cases likely represent the less severe end of the clinical spectrum. Medication effects were not characterized. Our findings should be interpreted as *proof-of-principle* that covariates can meaningfully confound biomarker-disorder associations, rather than as definitive claims about specific biomarkers in clinical populations. Third, cross-sectional biomarker measurements cannot establish temporal relationships; without longitudinal data, we cannot distinguish whether BMI and smoking are confounders, mediators, or colliders. Fourth, unmeasured factors—including diet and microbiome,^25–28^ sleep,^29^ medications and other substances like alcohol and caffeine,^25–27^, urbanicity^30^, menstrual cycle and menopause,^31,32^ and chronic diseases^33^—may explain residual associations. Fifth, analyses were restricted to biomarkers available in the UKB; other biomarkers of interest in psychiatric research (e.g., cortisol, oxytocin, melatonin) were not assessed, although the methodological principles described here are likely to generalize. Finally, our framework provides general guidance but cannot replace study-specific causal reasoning tailored to individual research questions and designs.

## Conclusions

Blood biomarkers show promise for psychiatric research but realizing their potential requires more rigorous and deliberate handling of covariates. Many biomarker-disorder associations are shaped by demographic, temporal, or behavioral factors that may go undetected in smaller studies. We emphasize careful study design, systematic covariate measurement (particularly time of day), sensitivity analyses, and transparent reporting across adjustment models. Longitudinal designs are essential to distinguish confounders from mediators. Adoption of these practices may improve reproducibility and accelerate identification of reliable and valid biomarkers that reflect disorder-specific pathophysiology rather than demographic or lifestyle confounding.

## Data Availability

All data produced in the present work are contained in the manuscript

## Acknowledgements

This work was supported by a National Health and Medical Research Council Synergy Grant (2019260), National Health and Medical Research Council Investigator Grants to J.J.C. (2008196), N.R.W. (1173790) and I.B.H. (2016346), Pioneer Centre for Statistical and computational Methods for Advanced Research to Transform Biomedicine (SMARTbiomed), DNRF grant number P4 (N.R.W.), the Michael Davys Trust (N.R.W.), a Lundbeck Foundation Postdoctoral Fellowship to C.A. (R449-2023-1077), and NARSAD Young Investigator Grants to C.A. (33624) and J.J.C. (32977).

