## Supplementary material for "Rethinking covariate adjustment in psychiatric biomarker research: a framework applied to UK Biobank blood samples": supp_material

**eMethods**

**Study population and data source**

This study utilized data from the UK Biobank (UKB), a large prospective cohort study that recruited approximately 500,000 participants aged 40-69 years between 2007 and 2010 across 22 assessment centers in the United Kingdom. The study design of the UKB has been described extensively.^1^ The UKB received approval from the Northwest Multicentre Research Ethics Committee (reference: 11/NW/0382), and this analysis was conducted under application number 116122. All participants provided written consent.

**Blood biomarker selection and platforms**

Detailed protocols for blood collection and handling have been reported previously.^2^ We identified 29 markers through a targeted literature review of blood-based markers that are frequently reported in psychiatric research and are available in the UKB (see Table S1). These biomarkers span four distinct analytical platforms. All markers were filtered to exclude participants having fasting time ≥ 24 hours, sample collection time outside 9:00-20:00 hours and with categorical variables n < 50.

*Biochemistry*: Biochemical assays were performed on serum samples using automated clinical chemistry analyzers at the UKB central laboratory. Values were log-transformed. We included 11 markers: Albumin, high sensitivity CRP, Creatinine, Cystatin C, Gamma-glutamyl transferase (GGT), Glucose, Glycated hemoglobin (HbA1c), Estradiol, Testosterone, Total bilirubin, and Total protein.

*Blood cell counts*: Complete blood counts were analyzed using Beckman Coulter LH750 Hematology Analyzers employing VCS (Volume, Conductivity, and Scatter) technology on 4ml EDTA samples. We excluded samples with time between venipuncture and processing exceeding 36 hours to maintain analytical quality. Values were log-transformed. We included four markers: Leukocyte count, Monocyte-to-Lymphocyte Ratio (MLR), Neutrophil-to-Lymphocyte Ratio (NLR), and Platelet-to-Lymphocyte Ratio (PLR).

*Metabolomics-NMR*: Nuclear magnetic resonance (NMR) spectroscopy was performed by Nightingale Health Ltd. on EDTA plasma samples. Samples were analyzed between June 2019 and June 2022. The platform measured 251 metabolic biomarkers from approximately 280,000 UKB participants. Values underwent log1p transformation, followed by standardization and rank-based inverse normal transformation (RINT). Standardized values exceeding 4 standard deviations were considered outliers and excluded from analysis, following established Nightingale protocols.^3^ We included four markers: Alanine, Low-density lipoprotein (LDL) cholesterol, Total Cholesterol, and Total Triglycerides.

*Proteomics-Olink:* Protein analysis was conducted using the Olink Explore 3072 platform on blood plasma samples. The platform measures 2,941 protein analytes capturing 2,923 unique proteins across eight 384-plex panels: Cardiometabolic I and II, Inflammation I and II, Neurology I and II, and Oncology I and II.^4^ Protein levels were reported as Normalized Protein eXpression (NPX) values on a log2 scale. Samples were processed between April 2021 and February 2022 from approximately 55,000 UKB participants. NPX values also underwent RINT. We included 10 markers: BDNF, Interleukin (IL)-6, IL-10, IL-12, IL-18, IL-1β, Leptin, Corticosteroid-binding globulin (Serpina 6), TNF-α, and Insulin-like growth factor-binding protein 1 (IGFBP1).

**Covariate classification**

Our primary objective was to quantify systematically the contribution of different potential covariates to biomarker variance. We classified covariates into four categories based on their nature and role in biomarker research.

*Technical factors:* We included assessment center identification (Field 54) and genetic principal components PC1-PC20 (Field 22009) as universal technical factors applied to all biomarkers. Genetic PCs capture biological ancestry variation rather than technical artefacts but are grouped here because they require universal adjustment to prevent population stratification bias in biomarker association studies. Biomarker levels were residualized for all technical covariates prior to downstream analyses. We also included self-reported fasting time (Field 74) as a technical factor, as it primarily reflects compliance with pre-collection protocols rather than biological fasting state. Platform-specific technical factors were incorporated based on known sources of analytical variation. For biochemistry, we included sample dilution factor (Field 30897) and assay date (Category 18518). For blood cell counts, we included device identification and processing time between sampling and analysis (Category 9081). For NMR metabolomics, processing batch (Field 20282), spectrometer identification (Field 23650), and time between sample preparation and measurement were included. For proteomics-Olink, we included processing batch, UKB-PPP consortium selection status (Field 30903), and panel processing dates.

*Standard demographic factors*: We included age at recruitment (Field 21002) and sex (Field 31) as established demographic factors known to influence biomarker concentrations across multiple platforms. Based on known hormonal transitions across the lifespan, we also examined age x sex interactions. Ancestry was additionally examined as a demographic factor in covariate-disorder association analyses. The influence of genetic ancestry on biomarker levels, before and after adjustment for genetic principal components, is characterized in Figures S3-S4.

*Temporal factors:* Time-related sources of biological variation included time of day at blood collection (Field 3166, continuous in hours) and month of assessment center attendance (Field 55, categorical 1-12) to capture diurnal and seasonal variation in biomarker levels. For disorder association analyses, months were grouped into seasons (winter: December-February; spring: March-May; summer: June-August; autumn: September-November).

*Behavioral factors*: We included body mass index (BMI, calculated as weight [Field 21001] divided by height squared [Field 50]), smoking status (Field 20116: never, previous, current), and chronotype (Field 1180: definitely morning, rather morning, don't know, rather evening, definitely evening).

**Psychiatric disorder definitions**

We used ICD-9 and ICD-10 codes in Hospital inpatient data (Category 2000), ICD-10 codes in Death Register records (Field 40001, Field 40002), and self-reported medical condition codes (Field 20002) reported at baseline or subsequent UK Biobank assessment center visits. Participants were classified as cases if they had any recorded diagnosis for the following: bipolar disorder (F31), depressive episode (F32), recurrent depression (F33), and schizophrenia (F20).

**Statistical analysis**

*Variance quantification analysis:* Analyses were conducted using the aov() function (R version 4.3.2) to quantify the proportion of variance in each biomarker explained by different covariate categories. The statistical model included all covariate categories simultaneously as following:

*biomarker ~ technical factors + age + sex + age x sex + BMI + month of assessment + time of day + smoking + chronotype*

For variance partitioning presentation, the variance explained by the sex × age interaction was equally allocated between sex and age categories. Partial R^2^ values were calculated as the ratio of sum of squares for each covariate term to total sum of squares, providing quantitative estimates of relative covariate importance.

*Temporal factor analysis:* We visualized selected biomarker levels (those for which linear regression effect was significant) across month of assessment (seasonal patterns) and time of day (diurnal patterns) to characterize temporal variation.

*Chronotype x study design comparison:* Data were drawn from two assessment waves of the UKB. During the initial assessment (UKB1), conducted between 2006 and 2010, all participants attended an assessment center where blood was collected. Participants received an invitation letter (<https://bb30.ndph.ox.ac.uk/ukb/ukb/docs/ac_invite.pdf>) with a provisional appointment date and time, which they were able to change if needed; however, the proportion of participants who rescheduled their appointments is unknown. Appointments at the initial assessment were scheduled to last approximately 2–3 hours. Several years later, a subset of participants was invited to take part in a repeat assessment (UKB2) as part of an imaging study (2014+). In contrast to the initial assessment, participants invited to the repeat assessment were required to actively schedule their appointment, allowing them to self-select a suitable date and time. Appointments at the repeat assessment were scheduled to last approximately 4-5 hours (<https://www.ukbiobank.ac.uk/wp-content/uploads/2025/01/Participant-information-sheet-initial-imaging-study.pdf>).

Blood sample time from 6,690 participants attending both UKB1 and UKB2 was analysed using a linear mixed-effects model (LMM) fitted with restricted maximum likelihood (REML) using the lme4 package (R version 4.3.2) in R, adding sex and age as fixed effects. Statistical inference for fixed effects was obtained using the lmerTest package, applying Satterthwaite’s approximation for degrees of freedom. The model included chronotype (five categories/levels) and UKB visit (UKB1 vs UKB2) as fixed effects, as well as their interaction, to test whether changes in blood sample time between visits differed by chronotype. To account for the repeated-measures structure of the data, participant ID was included as a random intercept. This modelling approach accounts for within-participant dependence while estimating differences in blood sample time across UKB visits and chronotype groups.

*Ancestry analysis:* To assess the influence of genetic ancestry on biomarker levels, we compared unadjusted and residualized (technical-adjusted) biomarker levels across six ancestry groups (Field 30079): European (EUR, reference), African (AFR), Admixed American (AMR), Central/South Asian (CSA), East Asian (EAS), and Middle Eastern (MID).

*Covariate-psychiatric disorder associations:* We tested associations between key covariates (sex, age, season, smoking, BMI, chronotype, time of day) and psychiatric disorder diagnoses using logistic regression. Results were corrected for multiple testing using false discovery rate (FDR) adjustment.

*Sequential adjustment analysis:* To evaluate the effects of relevant covariates on biomarker-disorder associations, we first adjusted all biomarker levels for technical covariates in a linear model: biomarker ~ technical factors and took the model’s residuals. We then tested associations between each residualized biomarker and psychiatric disorders under multiple adjustment levels: (1) unadjusted; (2) adjusted for individual covariates (time of day, age, sex, chronotype, season, smoking, BMI) separately; and (3) fully adjusted (all covariates jointly). We compared the proportion of significant associations retained across adjustment levels.

*Leukocyte count sensitivity analysis:* To examine whether biomarker-disorder associations were driven by general inflammatory processes, we tested correlations between each biomarker and leukocyte count, and compared biomarker-disorder odds ratios with and without additional adjustment for leukocyte count.

**Table S1. Summary of biomarkers in mood disorders** (based on review papers and available guidelines)

| **Metabolic Markers** | **In UKB?** | **UKB field ID** | **Final decision for the current paper** |
| --- | --- | --- | --- |
| **Glucose**: Recommended as baseline lab investigation for MDD and BD ^5,6^ | **Yes** | **20280:** Glucose-lactate (NMR)  **23470:** Glucose (NMR)  **30740:** Glucose (hexokinase analysis) | **30740:** Glucose (hexokinase analysis) |
| **HbA1c**: Shows bidirectional relationship with depression; elevated in MDD ^7,8^ | **Yes** | **30750:** Glycated haemoglobin (HbA1c) (HPLC analysis)  **30020:** Haemoglobin concentration ("Haemoglobin Concentration" assay) | **30750:** Glycated haemoglobin (HbA1c) (HPLC analysis) |
| **Insulin**: Increased in depression, particularly atypical depression; small increases in acute depression suggest it's a state biomarker rather than trait biomarker ^9,10^ | **Indirect** | See IGF-1 | — |
| **Adipokines** | | |  |
| **Leptin**: Increases with age and BMI more in BD than controls; higher in mild/moderate depression ^9,11^ | **Yes** | **LEPR:** OLINK (P48357) Cardiometabolic  **LEP**: OLINK (P41159) Cardiometabolic | **LEP**: OLINK (P41159) Cardiometabolic |
| **Adiponectin**: No overall differences between MDD and controls, but RIA assay method showed lower levels in MDD ^11^ | **Yes** | **NECTIN1:** OLINK (Q15223) Inflammation II  **NECTIN2:** OLINK (Q92692) Cardiometabolic  **NECTIN4:** OLINK (Q96NY8) Oncology | — |
| **Lipid Profile** | | |  |
| **Lipids**: Altered in depression; recommended as baseline test for MDD and BD ^5,6,10^ | **Yes** | Nightingale Health Biobank Collaborative Group. | **LDL cholesterol**  **Total cholesterol** |
| **Triglycerides**: High levels associated with perinatal depression ^12^ | **Yes** | Same as above | **Total triglycerides** |
| **HPA Axis** | | |  |
| **Cortisol**: Reduced in pregnant/postpartum women with depression; small but significant predictive effect on MDD onset/relapse; often elevated in depression ^10,13,14^ | **Indirect** | **SERPINA6**: OLINK (P08185) Inflammation II  **POMC**: OLINK (P01189) Cardiometabolic II  **HSD11B1**: OLINK (P28845) Inflammation | **SERPINA6** (CBG, Corticosteroid-binding globulin) |
| **Sex Hormones** | | |  |
| **Oestradiol**: Evidence suggests relationship with adolescent mood changes; higher during manic episodes ^15,16^ | **Yes** | **30800**: Oestradiol (two step competitive analysis) | **30800**: Oestradiol (two step competitive analysis) |
| **Testosterone**: Low in MDD; higher during manic vs. depressive episodes ^8,15^ | **Yes** | **30850**: Testosterone (one step competitive analysis) | **30850**: Testosterone (one step competitive analysis) |
| **Prolactin**: Evidence did not support changes as marker of MD during peripartum ^13^ | **Yes** | **PRL:** OLINK (P01236) Neurology | **PRL** |
| **Progesterone**: Higher during manic episodes ^15^ | **Indirect** | **PIBF1**: OLINK (Q8WXW3-4) Neurology II  **PGR**: OLINK (P06401) Inflammation II | — |
| **Thyroid Function** | | |  |
| **TSH**: Recommended for MDD with lithium and BD baseline; no evidence for changes during peripartum depression ^5,6,13^ | **Indirect** | **THRAP3:** OLINK (Q9Y2W1) Cardiometabolic II  **TSHB:** OLINK (P01222) Cardiometabolic  **PTH**: OLINK (P01270) Neurology II  **PTH1R**: OLINK (Q03431) Inflammation  **SERPINA7**: OLINK (P05543) Inflammation II  **TEF**: OLINK (Q10587) Cardiometabolic II  **TG**: OLINK (P01266) Oncology II | — |
| **Kidney Function Markers** | | |  |
| **Cystatin C**: High levels associated with male depression ^12^ | **Yes** | **30720**: Cystatin C (latex enhanced immunoturbidimetric analysis) | **30720**: Cystatin C |
| **Creatinine**: Recommended for MDD with lithium and BD baseline; low levels associated with male depression ^5,6,12^ | **Yes** | **23478**: Creatinine (NMR)  **30510**: Creatinine (enzymatic) in urine (enzymatic analysis)  **30700**: Creatinine (enzymatic analysis) | **30700**: Creatinine (enzymatic analysis) |
| **eGFR**: Recommended for MDD with lithium ^5^ | **Yes** | Function of Creatinine and Cystatin C | — |
| **Electrolytes** | | |  |
| **General Electrolytes**: Recommended for MDD with lithium and BD baseline ^5,6^ | **?** |  | — |
| **Magnesium**: Higher in patients on antidepressants/mood stabilizers; normal in untreated patients ^17^ | **No** |  | — |
| **Calcium**: Recommended as BD baseline test; low levels associated with female depression ^6,12^ | **Yes** | **30680**: Calcium (Arsenazo III analysis)  **SMOC1:**  **SMOC2:**  **CABP2:**  **NCS1:** | — |
| **Liver Function Markers** | | |  |
| **Albumin**: Lower levels associated with depression; low in MDD ^8,18^ | **Yes** | **23479**: Albumin (NMR)  **30600**: Albumin (BCG analysis) | **30600**: Albumin (BCG analysis) |
| **Total Protein**: Low levels strongly associated with female depression and perinatal depression ^12^ | **Yes** | **30860**: Total protein (biuret analysis) | **30860**: Total protein (biuret analysis) |
| **Bilirubin**: Recommended as BD baseline test; lower levels associated with depression ^6,18^ | **Yes** | **30660**: Direct bilirubin (DPD analysis)  **30840**: Total bilirubin (photometric colour analysis) | **30840**: Total bilirubin (photometric colour analysis) |
| **ALT**: Elevated in MDD ^8^ | **Yes** | **30620**: Alanine aminotransferase (IFCC analysis)  **20281**: Spectrometer-corrected alanine (NMR)  **23460**: Alanine (NMR)  **23468**: Phenylalanine (NMR) | **20281: Alanine (NMR)** |
| **General Liver Function**: Recommended at baseline and every 6-12 months for MDD with pre-existing liver disease and BD ^5,6^ | **?** |  | — |
| **AST**: Elevated in MDD ^8^ | **Yes** | **30650**: Aspartate aminotransferase (IFCC analysis)  **GOT1:** OLINK (P17174) Cardiometabolic II | — |
| **GGT**: Higher levels associated with depression; elevated in MDD ^8,18^ | **Yes** | **30730**: Gamma glutamyltransferase (IFCC analysis)  **TGM2:** OLINK (P21980) Cardiometabolic  **GGCT:** OLINK (O75223) Cardiometabolic II  **GGACT:** OLINK (Q9BVM4) Cardiometabolic II  **GGH:** OLINK (Q92820) Cardiometabolic  **CHAC2:** OLINK (Q8WUX2) Oncology  **23461**: Glutamine (NMR) | **30730**: Gamma glutamyltransferase (IFCC analysis) |
| **ALP**: Higher levels associated with depression ^18^ | **Yes** | **30610**: Alkaline phosphatase (AMP(IFCC) analysis)  **ALPI:** OLINK (P09923) Inflammation II  **ALPP:** OLINK (P05187) Oncology | — |
| **Neurotrophic Factors** | | |  |
| **BDNF**: Reduced in depression; decreased in mania and bipolar depression, but not in euthymia ^10,19^ | **Yes** | **BDNF:** OLINK (P23560) Cardiometabolic II | **BDNF** |
| **IGF-1**: Low levels associated with perinatal depression ^12^ | **Yes** | **IGFBP1:** OLINK P08833 Cardiometabolic | **IGFBP** |
| **VEGF**: Elevated in depression ^10^ | **Yes** | **VEGFA:**  **VEGFB:**  **VEGFC:**  **VEGFD:**  **FLT1:**  **FLT3:**  **FLT4:**  **KDR:** | — |
| **Inflammatory Markers** | | |  |
| **CRP**: Bidirectional relationship with depressive symptoms; elevated in depression (particularly atypical), euthymia, and mania; higher during manic than depressive episodes ^10,15,18-21^ | **Yes** | **30710**: C-reactive protein (immunoturbidimetric - high sensitivity analysis) | **30710**: C-reactive protein |
| **Cytokines** | | |  |
| **TNF-α**: Increased in acute schizophrenia, bipolar mania, and MDD; elevated in MDD compared to controls; increased in mania and bipolar depression, but not in euthymia ^10,19,20,22,23^ | **Yes** | **TNF:** OLINK P01375 Cardiometabolic | **TNF** |
| **IL-6**: Increased in acute schizophrenia, MDD, euthymic bipolar disorder; elevated in MDD; consistent association linking depression with future IL-6; increased in mania and euthymia ^10,19-23^ | **Yes** | **IL6:** OLINK P05231 Oncology | **IL6** |
| **IL-1β**: Increased in chronic schizophrenia and euthymic bipolar disorder ^22^ | **Yes** | **IL1B:** OLINK P01584 Inflammation | **IL1B** |
| **IL-12**: Elevated in MDD compared to controls ^23^ | **Yes** | **IL12A_IL12B:** OLINK P29459_P29460 Oncology | **IL12A_IL12B** |
| **IL-18**: Elevated in MDD compared to controls ^23^ | **Yes** | **IL18:** OLINK Q14116 Inflammation | **IL18** |
| **IL-10**: Elevated in MDD compared to controls ^23^ | **Yes** | **IL10:** OLINK P22301 Inflammation | **IL10** |
| **IL-13**: Elevated in MDD compared to controls ^23^ | **Yes** | **IL13:**  **IL13RA1:**  **IL13RA2:** | — |
| **IFN-γ**: Slightly reduced in MDD compared to controls ^23^ | **Yes** | **IFNG:**  **IFNGR1:**  **IFNGR2:** | — |
| **White Blood Cells** | | |  |
| **WBC Count**: Increased in depression; overall leukocytosis in depression; increased in MDD ^8,24,25^ | **Yes** | **3000: White blood cell (leukocyte) count** | **3000: Leukocyte counts** |
| **Lymphocytes**: Decreased in depression ^25^ | **Yes** |  | — |
| **Granulocytes**: Increased mean in depression ^24^ | **Yes** |  | — |
| **Neutrophils**: Increased in depression; higher levels associated with depression; increased in MDD ^8,18,24^ | **Yes** |  | — |
| **Monocytes**: Increased mean in depression; lower percentage associated with depression ^18,24^ | **Yes** |  | — |
| **CD4+ helper T cells**: Increased mean in depression ^24^ | **Yes** |  | — |
| **Natural killer cells**: Increased mean in depression; reduced NK cell cytotoxicity in depression ^24,25^ | **Yes** |  | — |
| **B cells**: Increased mean in depression ^24^ | **Yes** |  | — |
| **Activated T cells**: Increased mean in depression ^24^ | **Yes** |  | — |
| **Basophils**: Higher absolute counts associated with depression ^18^ | **Yes** |  | — |
| **CD4/CD8 ratio**: Increased in depression ^25^ | **Yes** |  | — |
| **Red Blood Cells** | | |  |
| **RBC Count**: Low count (anemia indicator) strongly associated with incident MDD ^8^ | **Yes** |  | — |
| **Haemoglobin**: Low concentration associated with incident MDD ^8^ | **Yes** |  | — |
| **Haematocrit**: Low percentage associated with incident MDD ^8^ | **Yes** |  | — |
| **Mean Corpuscular Volume (MCV)**: High MCV (macrocytosis indicator) associated with increased MDD incidence ^8^ | **Yes** |  | — |
| **Inflammatory Ratios** | | |  |
| **Neutrophil-to-Lymphocyte Ratio (NLR)**: Significantly higher in depressed patients; higher in BD compared to controls ^26,27^ | **Yes** | **30140: Neutrophil count**  **/**  **30120: Lymphocyte count** | **NLR** |
| **Platelet-to-Lymphocyte Ratio (PLR)**: Significantly higher in depressed patients; higher in BD compared to controls ^26,27^ | **Yes** | **30140: Platelet count**  **/**  **30120: Lymphocyte count** | **PLR** |
| **Monocyte-to-Lymphocyte Ratio (MLR)**: Slightly higher in depressed individuals ^26^ | **Yes** | **30130: Monocyte count**  **/**  **30120: Lymphocyte count** | **MLR** |
| **Coagulation** | | |  |
| **Prothrombin time and partial thromboplastin time**: Recommended as BD baseline test ^6^ | **Yes** | **F2: Not found**  **F2R:** OLINK (P25116) Inflammation | — |
| **Others** | | |  |
| **Vitamin D**: Low concentration associated with depression; associated with both female and male depression ^12,28^ | **Yes** | **30890**: Vitamin D (CLIA analysis) | — |

**eResults**

**Table S2.** Demographic and behavioral characteristics by psychiatric disorder status.

|  | **Bipolar disorder (N=1686)** | **Schizophrenia (N=855)** | **Recurrent depression (N=2621)** | **Depressive episode (N=40417)** | **Control (N=425995)** | **Statistic** | **P-value** |
| --- | --- | --- | --- | --- | --- | --- | --- |
| **Time of day** |  |  |  |  |  |  |  |
| 09:00-12:00 | 359 (21.3%) | 132 (15.4%) | 571 (21.8%) | 9366 (23.2%) | 105944 (24.9%) | 144.02 | 9.21e-25 |
| 12:00-15:00 | 530 (31.4%) | 271 (31.7%) | 830 (31.7%) | 12860 (31.8%) | 130816 (30.7%) |  |  |
| 15:00-18:00 | 558 (33.1%) | 314 (36.7%) | 875 (33.4%) | 12603 (31.2%) | 128987 (30.3%) |  |  |
| 18:00-21:00 | 234 (13.9%) | 136 (15.9%) | 336 (12.8%) | 5477 (13.6%) | 58545 (13.7%) |  |  |
| Missing | 5 (0.3%) | 2 (0.2%) | 9 (0.3%) | 111 (0.3%) | 1703 (0.4%) |  |  |
| **Genetically-inferred ancestry** |  |  |  |  |  |  |  |
| European ancestry (EUR) | 1427 (84.6%) | 650 (76.0%) | 2290 (87.4%) | 35601 (88.1%) | 367405 (86.2%) | 552.67 | 2.60e-104 |
| Admixed American ancestry (AMR) | 5 (0.3%) | 3 (0.4%) | 4 (0.2%) | 78 (0.2%) | 831 (0.2%) |  |  |
| African ancestry (AFR) | 24 (1.4%) | 46 (5.4%) | 22 (0.8%) | 258 (0.6%) | 6013 (1.4%) |  |  |
| Central/South Asian ancestry (CSA) | 22 (1.3%) | 31 (3.6%) | 35 (1.3%) | 464 (1.1%) | 7816 (1.8%) |  |  |
| East Asian ancestry (EAS) | 2 (0.1%) | 2 (0.2%) | 5 (0.2%) | 77 (0.2%) | 2534 (0.6%) |  |  |
| Middle Eastern ancestry (MID) | 4 (0.2%) | 6 (0.7%) | 6 (0.2%) | 144 (0.4%) | 1341 (0.3%) |  |  |
| Missing | 202 (12.0%) | 117 (13.7%) | 259 (9.9%) | 3795 (9.4%) | 40055 (9.4%) |  |  |
| **Age group** |  |  |  |  |  |  |  |
| 40–44 | 197 (11.7%) | 133 (15.6%) | 279 (10.6%) | 4448 (11.0%) | 43272 (10.2%) | 1128.62 | 1.36e-226 |
| 45–49 | 268 (15.9%) | 156 (18.2%) | 395 (15.1%) | 6050 (15.0%) | 54987 (12.9%) |  |  |
| 50–54 | 279 (16.5%) | 145 (17.0%) | 447 (17.1%) | 6992 (17.3%) | 63711 (15.0%) |  |  |
| 55–59 | 331 (19.6%) | 157 (18.4%) | 502 (19.2%) | 8087 (20.0%) | 76589 (18.0%) |  |  |
| 60–64 | 372 (22.1%) | 153 (17.9%) | 616 (23.5%) | 9129 (22.6%) | 104282 (24.5%) |  |  |
| 65–69 | 232 (13.8%) | 108 (12.6%) | 369 (14.1%) | 5564 (13.8%) | 81077 (19.0%) |  |  |
| Missing | 7 (0.4%) | 3 (0.4%) | 13 (0.5%) | 147 (0.4%) | 2077 (0.5%) |  |  |
| **Sex** |  |  |  |  |  |  |  |
| Female | 956 (56.7%) | 294 (34.4%) | 1733 (66.1%) | 26475 (65.5%) | 225600 (53.0%) | 2631.32 | <1e-300 |
| Male | 730 (43.3%) | 561 (65.6%) | 888 (33.9%) | 13942 (34.5%) | 200395 (47.0%) |  |  |
| **Chronotype** |  |  |  |  |  |  |  |
| Definitely morning | 361 (21.4%) | 154 (18.0%) | 585 (22.3%) | 8355 (20.7%) | 104642 (24.6%) | 2072.05 | <1e-300 |
| Rather morning | 395 (23.4%) | 172 (20.1%) | 632 (24.1%) | 11396 (28.2%) | 137556 (32.3%) |  |  |
| Don't know | 186 (11.0%) | 125 (14.6%) | 267 (10.2%) | 3979 (9.8%) | 44884 (10.5%) |  |  |
| Rather evening | 488 (28.9%) | 271 (31.7%) | 781 (29.8%) | 11847 (29.3%) | 106633 (25.0%) |  |  |
| Definitely evening | 256 (15.2%) | 133 (15.6%) | 356 (13.6%) | 4840 (12.0%) | 32280 (7.6%) |  |  |
| **Season** |  |  |  |  |  |  |  |
| Winter | 367 (21.8%) | 185 (21.6%) | 509 (19.4%) | 8393 (20.8%) | 89609 (21.0%) | 35.29 | 4.21e-04 |
| Fall | 395 (23.4%) | 186 (21.8%) | 603 (23.0%) | 9473 (23.4%) | 102229 (24.0%) |  |  |
| Spring | 481 (28.5%) | 241 (28.2%) | 811 (30.9%) | 12058 (29.8%) | 122400 (28.7%) |  |  |
| Summer | 443 (26.3%) | 243 (28.4%) | 698 (26.6%) | 10493 (26.0%) | 111757 (26.2%) |  |  |
| **Smoking status** |  |  |  |  |  |  |  |
| Never | 747 (44.3%) | 315 (36.8%) | 1283 (49.0%) | 19603 (48.5%) | 237667 (55.8%) | 3015.12 | <1e-300 |
| Previous | 535 (31.7%) | 241 (28.2%) | 856 (32.7%) | 14248 (35.3%) | 147265 (34.6%) |  |  |
| Current | 404 (24.0%) | 299 (35.0%) | 482 (18.4%) | 6566 (16.2%) | 41063 (9.6%) |  |  |
| **BMI group** |  |  |  |  |  |  |  |
| Healthy weight (18.5–24.9) | 445 (26.4%) | 229 (26.8%) | 681 (26.0%) | 11438 (28.3%) | 141203 (33.1%) | 2109.61 | <1e-300 |
| Underweight (<18.5) | 7 (0.4%) | 8 (0.9%) | 27 (1.0%) | 230 (0.6%) | 2141 (0.5%) |  |  |
| Overweight (25–29.9) | 632 (37.5%) | 298 (34.9%) | 1033 (39.4%) | 16033 (39.7%) | 182066 (42.7%) |  |  |
| Obese (30–39.9) | 513 (30.4%) | 268 (31.3%) | 756 (28.8%) | 11032 (27.3%) | 91960 (21.6%) |  |  |
| Severely obese (≥40) | 76 (4.5%) | 44 (5.1%) | 104 (4.0%) | 1474 (3.6%) | 7064 (1.7%) |  |  |
| Missing | 13 (0.8%) | 8 (0.9%) | 20 (0.8%) | 210 (0.5%) | 1561 (0.4%) |  |  |

**Figure S1. Variance in measurements of 29 biomarkers explained by technical, demographic, temporal, and behavioral factors.**

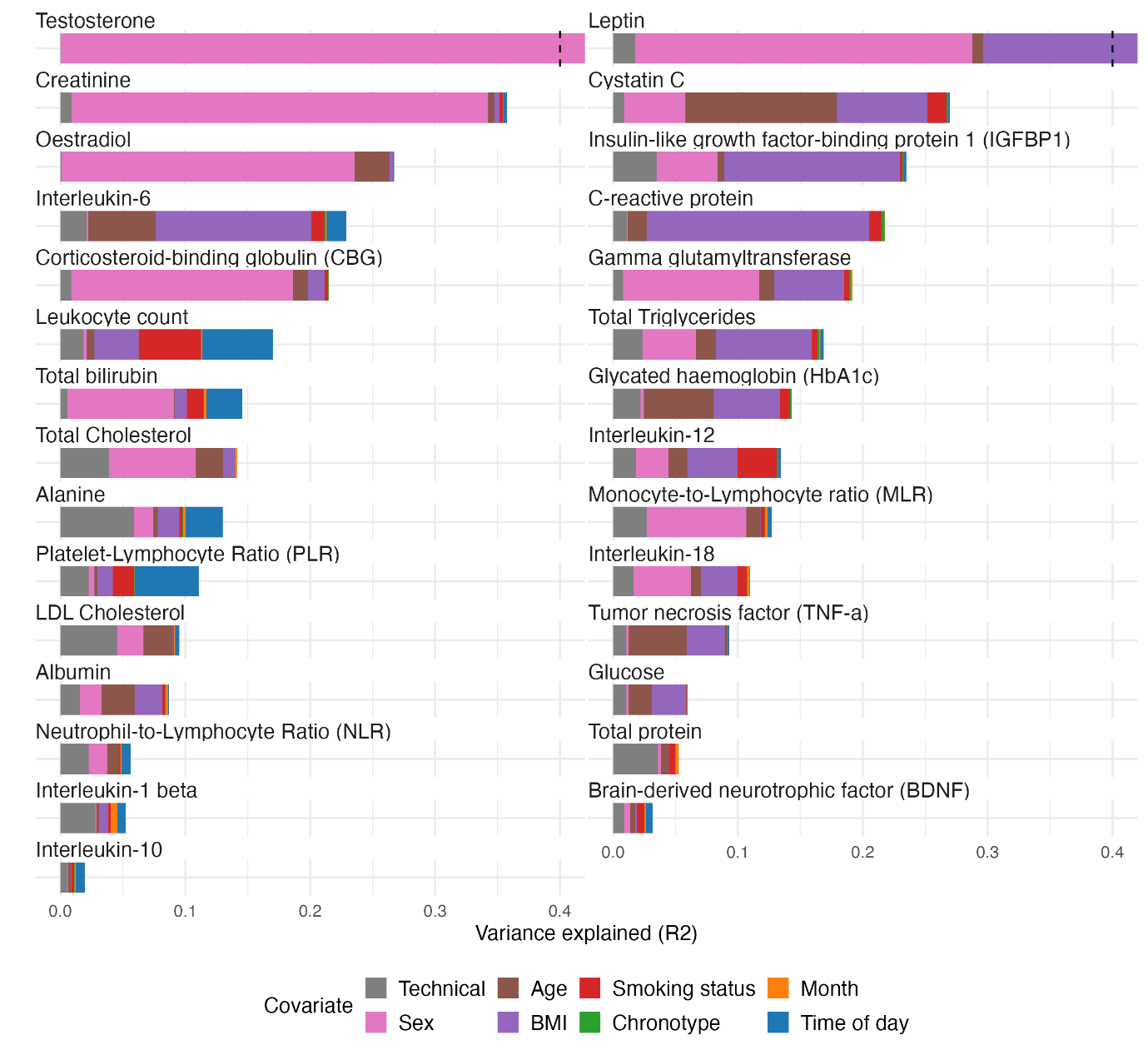

**Note**: the dotted line in the bar for testosterone and leptin indicates that the total variance explained was > 0.4 R^2^.

**Supplementary Figure S2.** Detailed breakdown of technical factor contributions to biomarker variance.

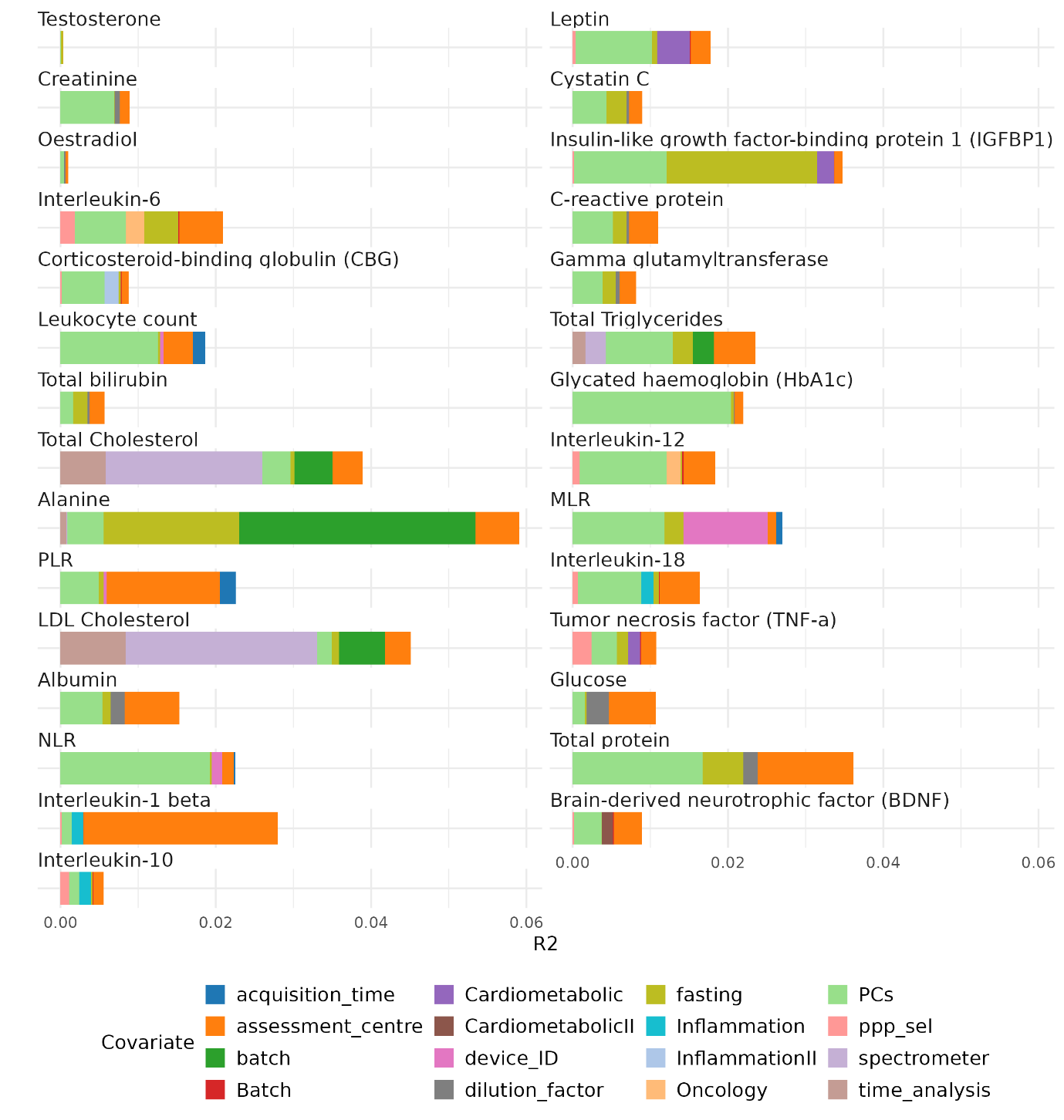

**Figure S3. Age-by-sex interaction effects on 29 biomarker levels after adjustment for technical covariates.**

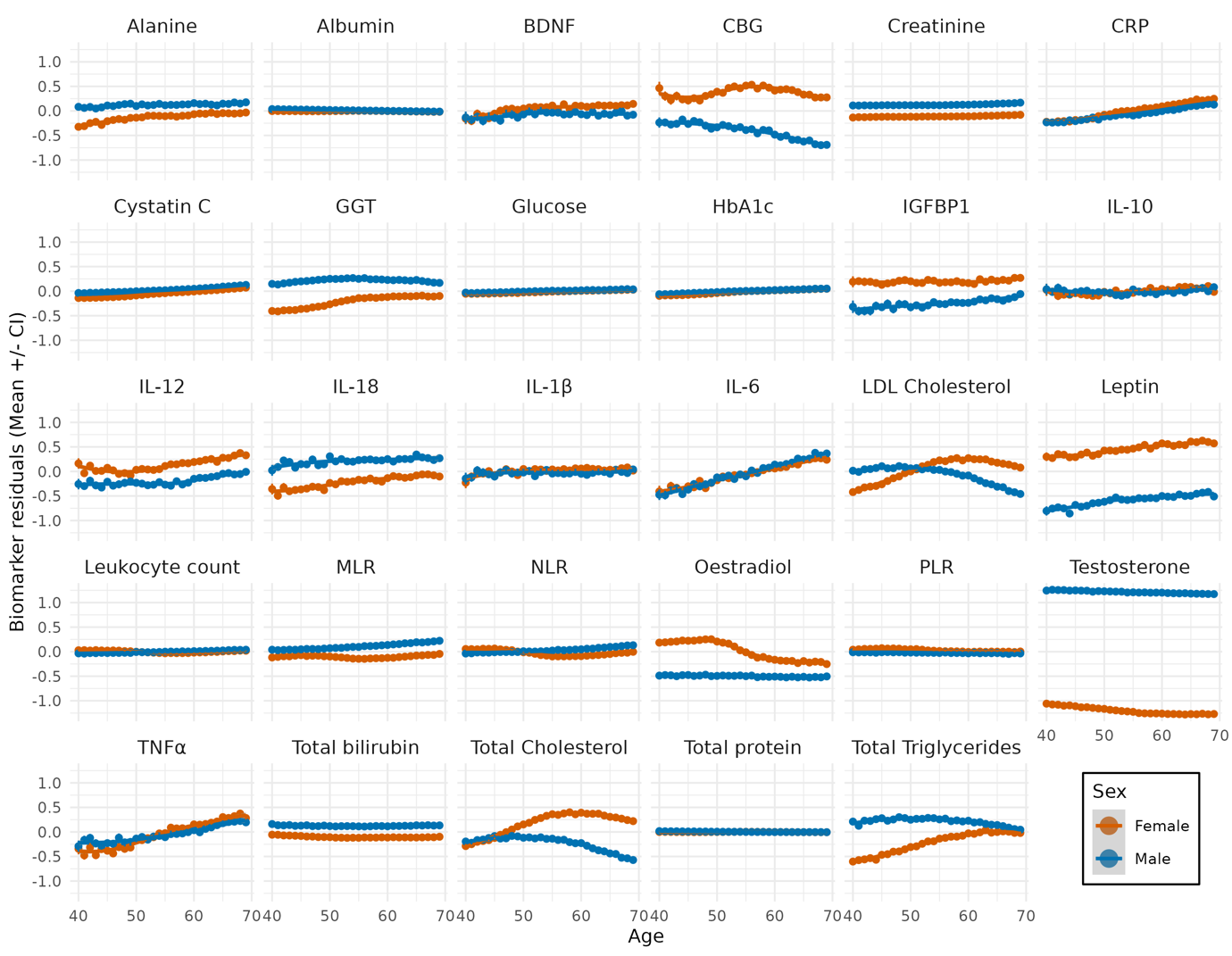

**Note:** After adjusting for technical covariates, pronounced age-by-sex interactions were observed across multiple biomarkers, highlighting that age-related changes differ substantially between sexes. Testosterone was consistently higher in males irrespective of age. Estradiol was higher in females at younger ages but declined sharply after age 50, coinciding with menopause, whereas levels in male remained stable across ages. LDL cholesterol and total cholesterol showed *crossover* effects around age 50-55, with males having higher levels at younger ages, but female levels increased sharply during menopause, surpassing male levels at older ages. Leptin was consistently higher in females than males. IL-6 and TNF-α increased with age in both sexes without clear sex differences. Several biomarkers including albumin, creatinine, cystatin C, glucose, HbA1c, and total protein showed minimal variance across age and sex after technical adjustment.

**Figure S4.** Unadjusted biomarker levels by genetic ancestry

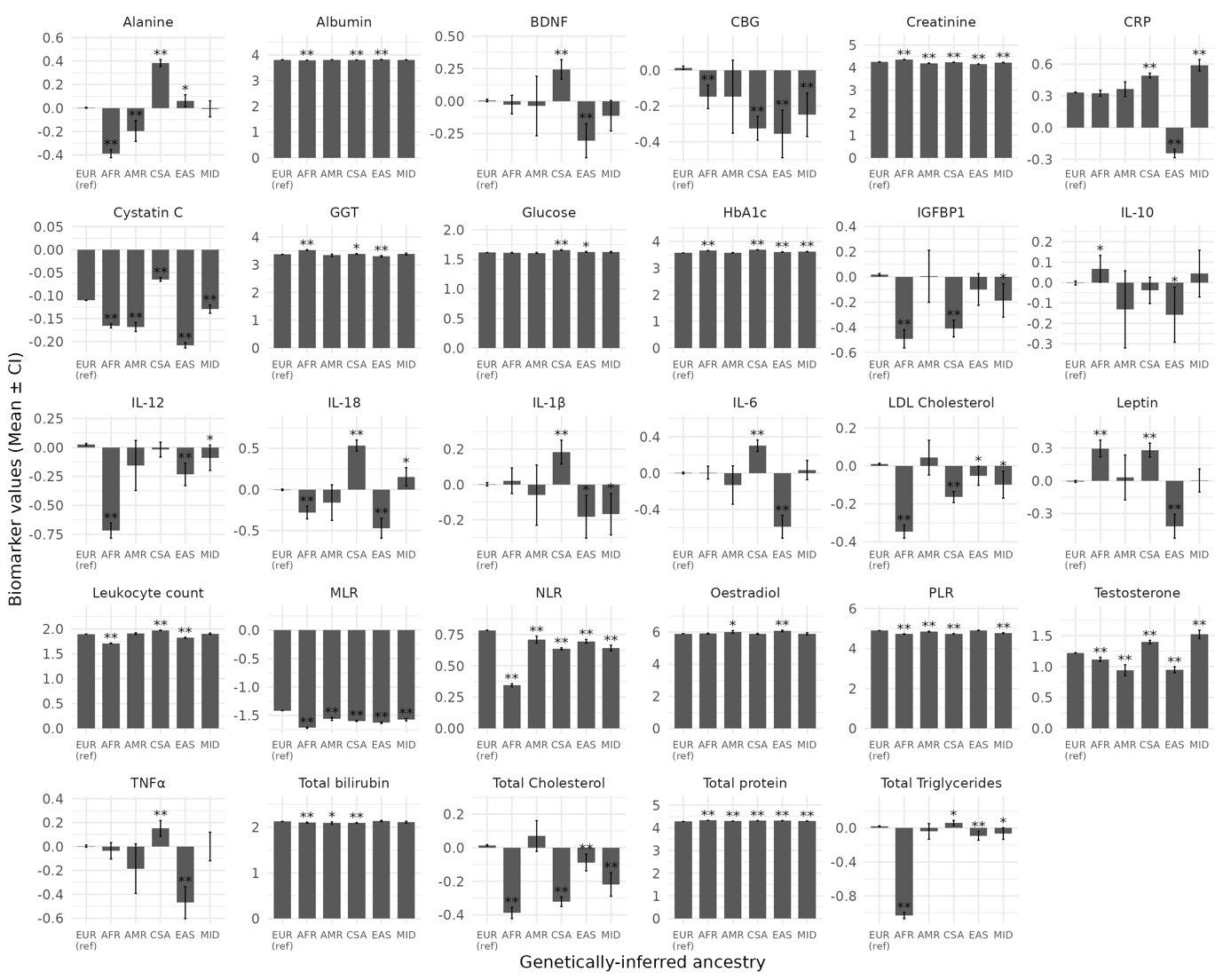

**Note:** Mean unadjusted biomarker levels across six genetic ancestry groups: European (EUR, reference), African (AFR), Admixed American (AMR), Central/South Asian (CSA), East Asian (EAS), and Middle Eastern (MID). Values are shown as differences from the EUR reference group. Error bars represent 95% confidence intervals. Ancestry categories were derived from UKB genetic ancestry classification (Field 30079). *p < 0.05; **FDR < 0.05.

**Figure S5.** Residualized biomarker levels by genetic ancestry after adjustment for genetic principal components

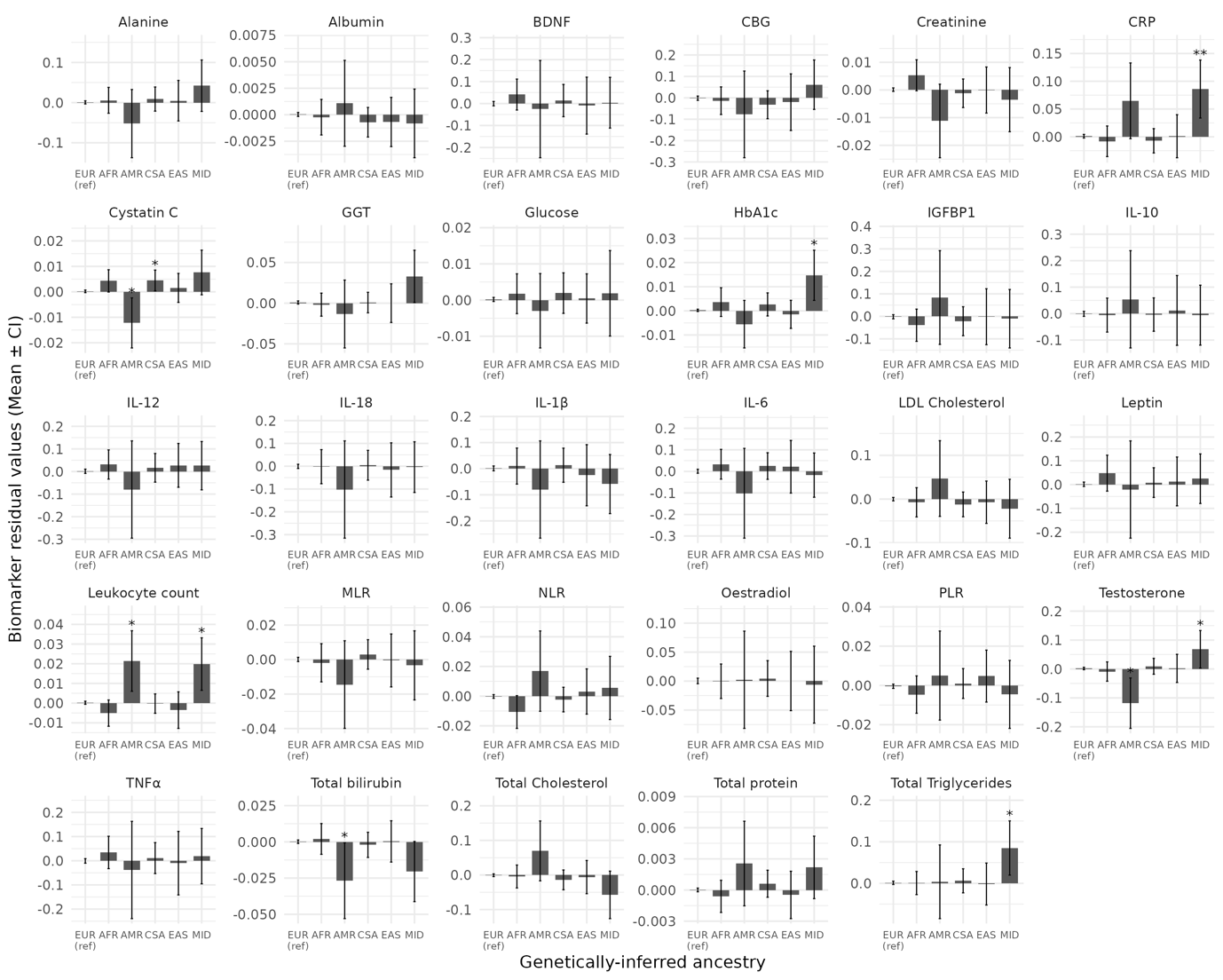

**Note:** Mean biomarker residual values after adjustment for technical covariates including 20 genetic principal components, shown across six genetic ancestry groups: European (EUR, reference), African (AFR), Admixed American (AMR), Central/South Asian (CSA), East Asian (EAS), and Middle Eastern (MID). Values are shown as differences from the EUR reference group. Error bars represent 95% confidence intervals. Most ancestry-related differences observed in unadjusted values (Figure S3) were attenuated after adjustment, with only CRP in the Middle Eastern group retaining significance after FDR correction. *p < 0.05; **FDR < 0.05.

**Figure S6.** Blood collection times of UKB across initial and imaging visits by self-reported chronotype.

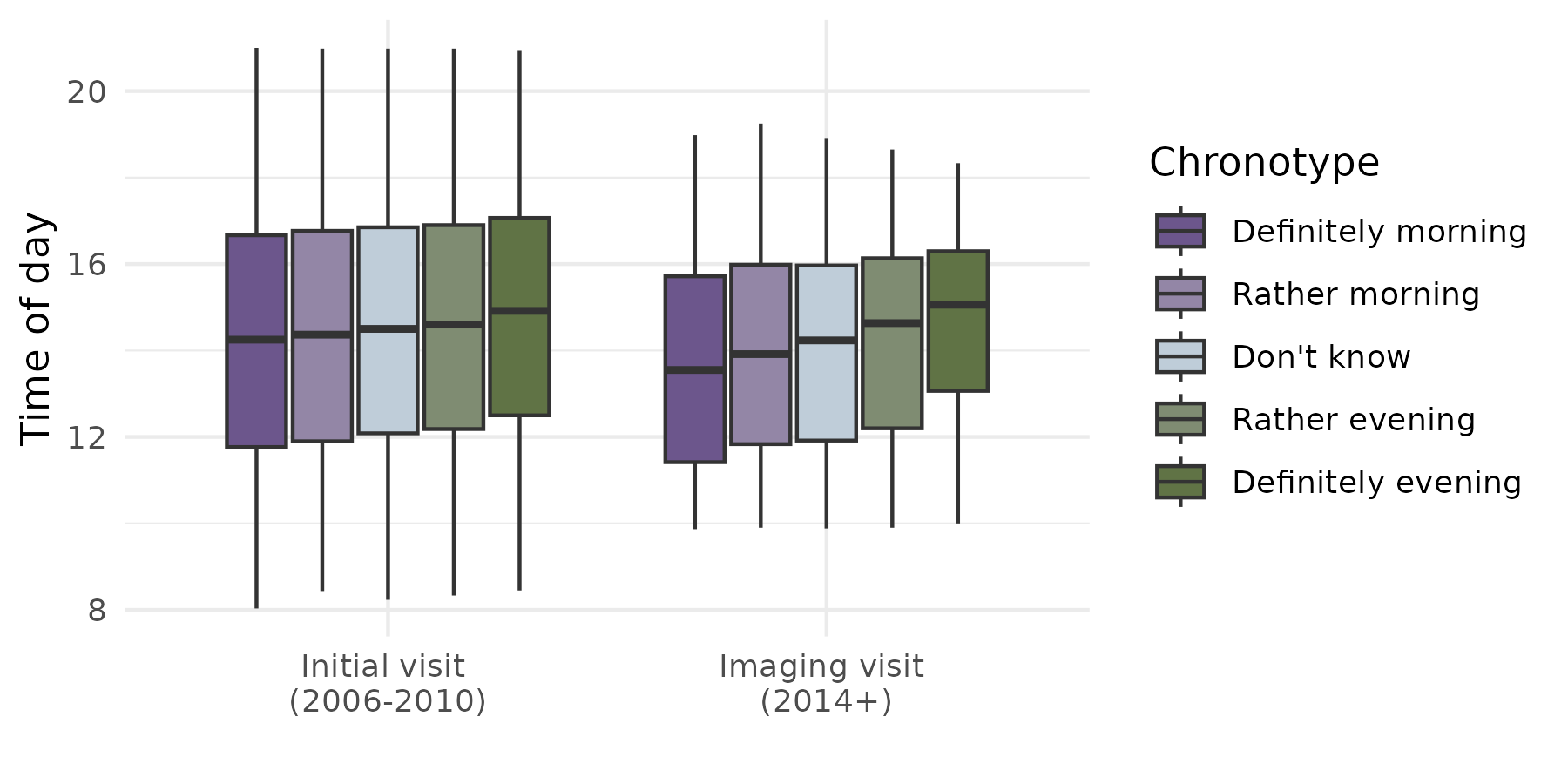

**Note:** Boxplots show the distribution of blood collection time (hour of day) across five chronotype categories at the initial assessment visit (UKB1, 2006-2010) and the repeat imaging visit (UKB2, 2014+). At UKB2, where participants self-scheduled their appointments, a clearer gradient by chronotype is visible compared to UKB1, where appointment times were pre-assigned.

**Figure S7.** Sequential covariate adjustment effects on biomarker-disorder associations for all 29 biomarkers across four psychiatric disorder categories.

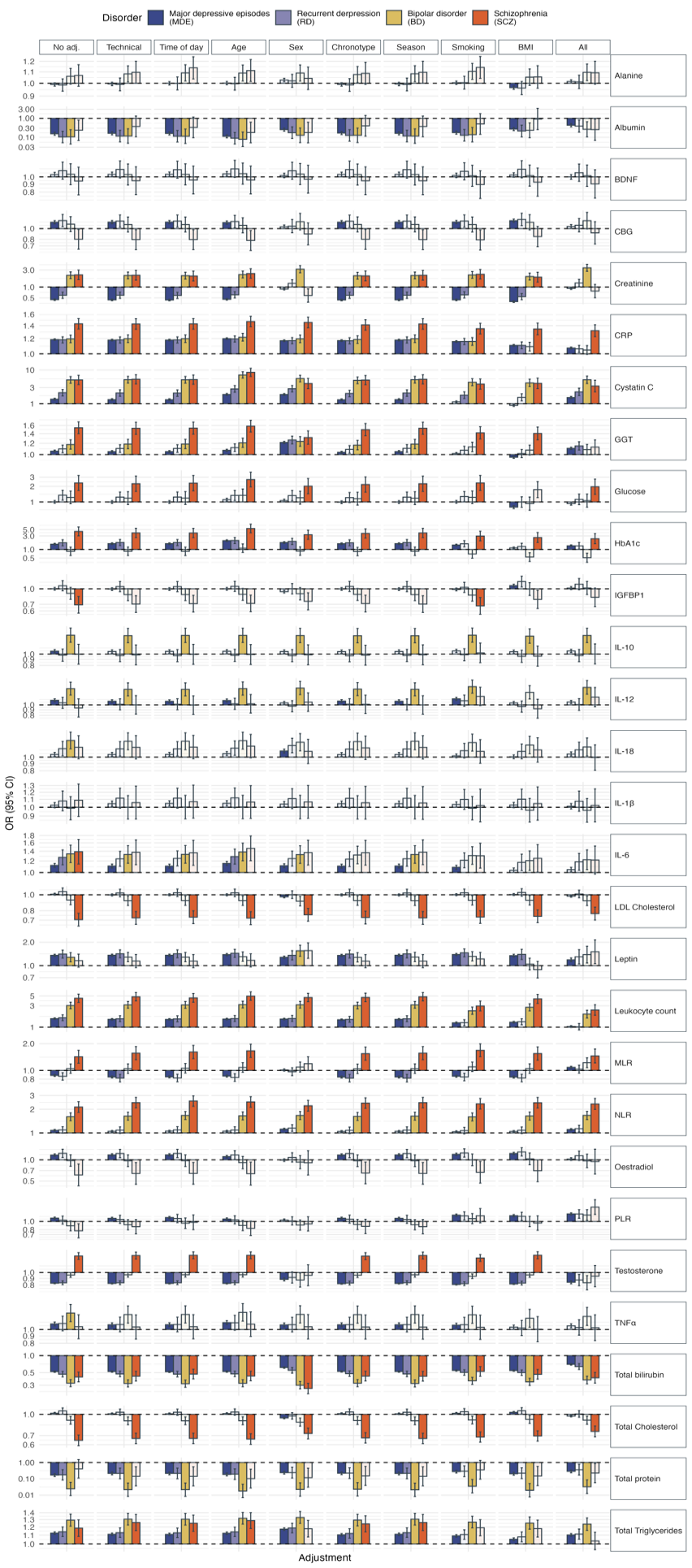

Note: All associations with single covariate (excluding the unadjusted model) were performed on the residuals of the biomarker levels after adjusting for technical covariates.

**Figure S8.** Leukocyte count associations
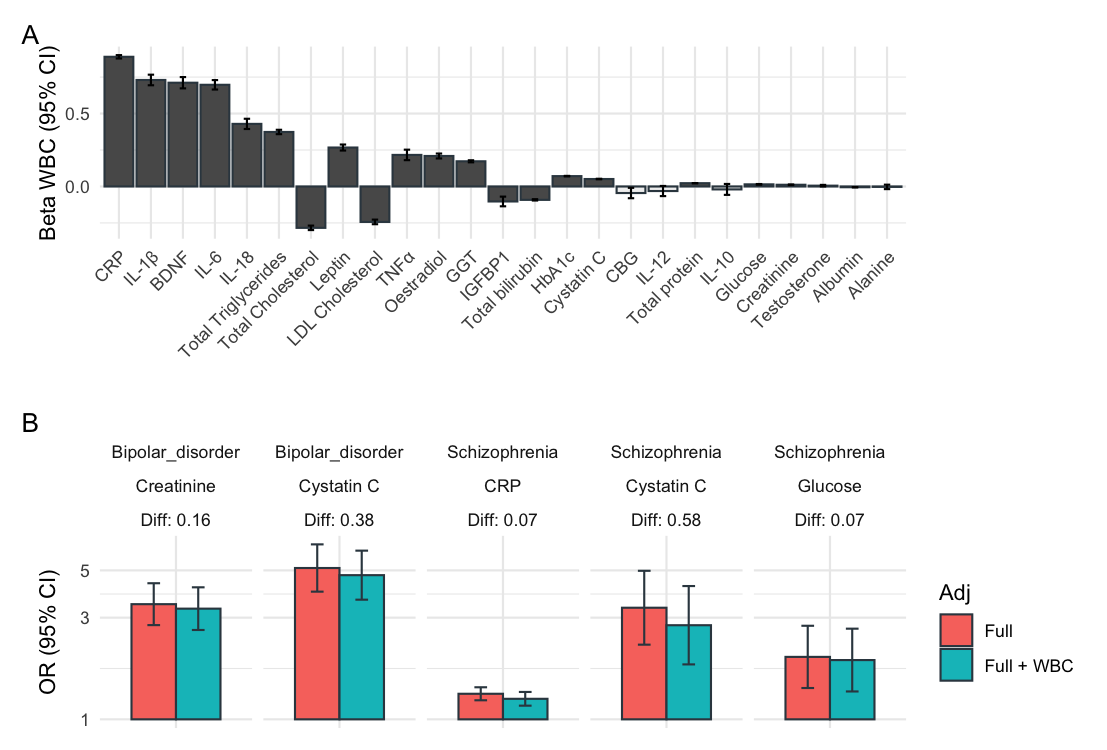

**Note:** Panel A: Association between leukocyte count and each biomarker (effect sizes from model: biomarker ~ WBC + covariates), ordered by effect size. Panel B: Comparison of biomarker-disorder ORs with and without leukocyte count adjustment, restricted to pairs showing a meaningful difference (for pairs with |ΔOR| > 0.05). Diff = absolute difference in OR between the two adjustment models.
